# SARS-CoV-2 vaccine breakthrough infections with the alpha variant are asymptomatic or mildly symptomatic among health care workers

**DOI:** 10.1101/2021.06.29.21259500

**Authors:** Francesca Rovida, Irene Cassaniti, Stefania Paolucci, Elena Percivalle, Antonella Sarasini, Antonio Piralla, Federica Giardina, Jose Camilla Sammartino, Alessandro Ferrari, Federica Bergami, Alba Muzzi, Viola Novelli, Alessandro Meloni, Sara Cutti, Anna Maria Grugnetti, Giuseppina Grugnetti, Claudia Rona, Marinella Daglio, Carlo Marena, Antonio Triarico, Daniele Lilleri, Fausto Baldanti

**Affiliations:** Molecular Virology Unit, Microbiology and Virology Department, Fondazione IRCCS Policlinico San Matteo, Pavia, Italy; Department of Clinical, Surgical, Diagnostic and Pediatric Sciences, University of Pavia, Pavia, Italy; Medical Direction, Fondazione IRCCS Policlinico San Matteo, Pavia, Italy; Department of Public Health, Experimental and Forensic Medicine, Section of Hygiene, University of Pavia, Pavia, Italy; Health Professions Direction, Fondazione IRCCS Policlinico San Matteo, Pavia, Italy; Direzione Sanitaria, Fondazione IRCCS Policlinico San Matteo, Pavia, Italy

## Abstract

Vaccine breakthrough SARS-CoV-2 infection has been monitored in 3720 healthcare workers receiving 2 doses of BNT162b2. SARS-CoV-2 infection is detected in 33 subjects, with a 100-day cumulative incidence of 0.93%. Vaccine protection against acquisition of SARS-CoV-2 infection is 83% (95%CI: 58-93%) in the overall population and 93% (95%CI: 69-99%) in SARS-CoV-2-experienced subjects, when compared with a non-vaccinated control group from the same Institution, in which SARS-CoV-2 infection occurs in 20/346 subjects (100-day cumulative incidence: 5.78%). The infection is symptomatic in 16 (48%) vaccinated subjects vs 17 (85%) controls (p=0.001). All analyzed patients, in whom the amount of viral RNA was sufficient for genome sequencing, results infected by the alpha variant. Antibody and T-cell responses are not reduced in subjects with breakthrough infection. Evidence of virus transmission, determined by contact tracing, is observed in two (6.1%) cases. This real-world data support the protective effect of BNT162b2 vaccine. A triple antigenic exposure, such as two-dose vaccine schedule in experienced subjects, may confer a higher protection.

## Introduction

Since the identification of severe acute respiratory syndrome coronavirus 2 (SARS-CoV-2) as etiological agent of Coronavirus Disease 19 (COVID-19), several efforts have been made in order to prevent infection and disease. Moreover, recently, highly effective vaccines have been introduced [1–4].

The licensed vaccines showed high efficacy in protection from SARS-CoV-2 infection in clinical trials, ranging from 70 to 95% [1–4]. However, post-authorization real-life studies are an important complement to evaluate the vaccine effectiveness in different populations and in the face of non-controlled real world challenges.

Initial nationwide data collection are confirming the effectiveness of the licensed vaccines, showing an effect size consistent with that reported in clinical trials [5–8].

However, clinical, virological and immunological characteristics of breakthrough SARS-CoV-2 infections after vaccination have been poorly investigated, due to lack of prospective systematical testing in vaccinated cohorts. Some studies conducted on healthcare workers reported a lower rate of symptomatic vs asymptomatic infections in vaccinated with respect to unvaccinated individuals [9–13], while data on the actual presence of infectious virus in SARS-CoV-2 RNA-positive samples recovered from vaccinated individuals are missing. Whether infected vaccinated subjects can transmit the infection, and to which extent, is a major concern for public health policy. Finally, whether post vaccine infections are associated with a deficient immune response to vaccination has not been investigated yet.

Healthcare workers have a high risk of exposure to SARS-CoV-2, therefore representing a challenging cohort for the evaluation of vaccine effectiveness and breakthrough infections. In Italy, the vaccination campaign started on December 27^th^, 2020, prioritizing healthcare workers and fragile and elderly individuals [14].

Aim of the present study is to investigate prospectively the risk of SARS-CoV-2 infection in vaccinated healthcare workers in a single Italian Center (Fondazione IRCCS Policlinico San Matteo, Pavia). Data are compared with the group of healthcare workers of the same Institution that did not receive the vaccination during the study period.

The characteristics of breakthrough infections, the underlying immune response and the risk of virus transmission to other individuals are investigated.

Results show that BNT162b2 vaccine is effective in reducing the incidence of SARS-CoV-2 infection in healthcare workers, while breakthrough infections are poorly symptomatic and are infrequently transmitted.

## Results

### Incidence and clinical characteristics of SARS-CoV-2 infection in vaccinated and non-vaccinated healthcare workers

During the period January 18-March 31 2021, 3720 healthcare workers received the second dose of BNT162b2 vaccine. Overall, before vaccination 507 subjects resulted SARS-CoV-2-experienced and 2761 SARS-CoV-2-naïve, while SARS-CoV-2 serostatus was unknown for 426 subjects and dubious for 26 subjects (Fig 1). After complete vaccination schedule, SARS-CoV2 infection was detected in 33 subjects (median time: 47, range 7-90, days after vaccination): 2 subjects among the 507 SARS-CoV-2-experienced, 24 among the 2761 SARS-CoV-2 naïve individuals, and 7 among the 452 individuals with unknown or dubious serostatus (Fig 1).

**Figure 1.**
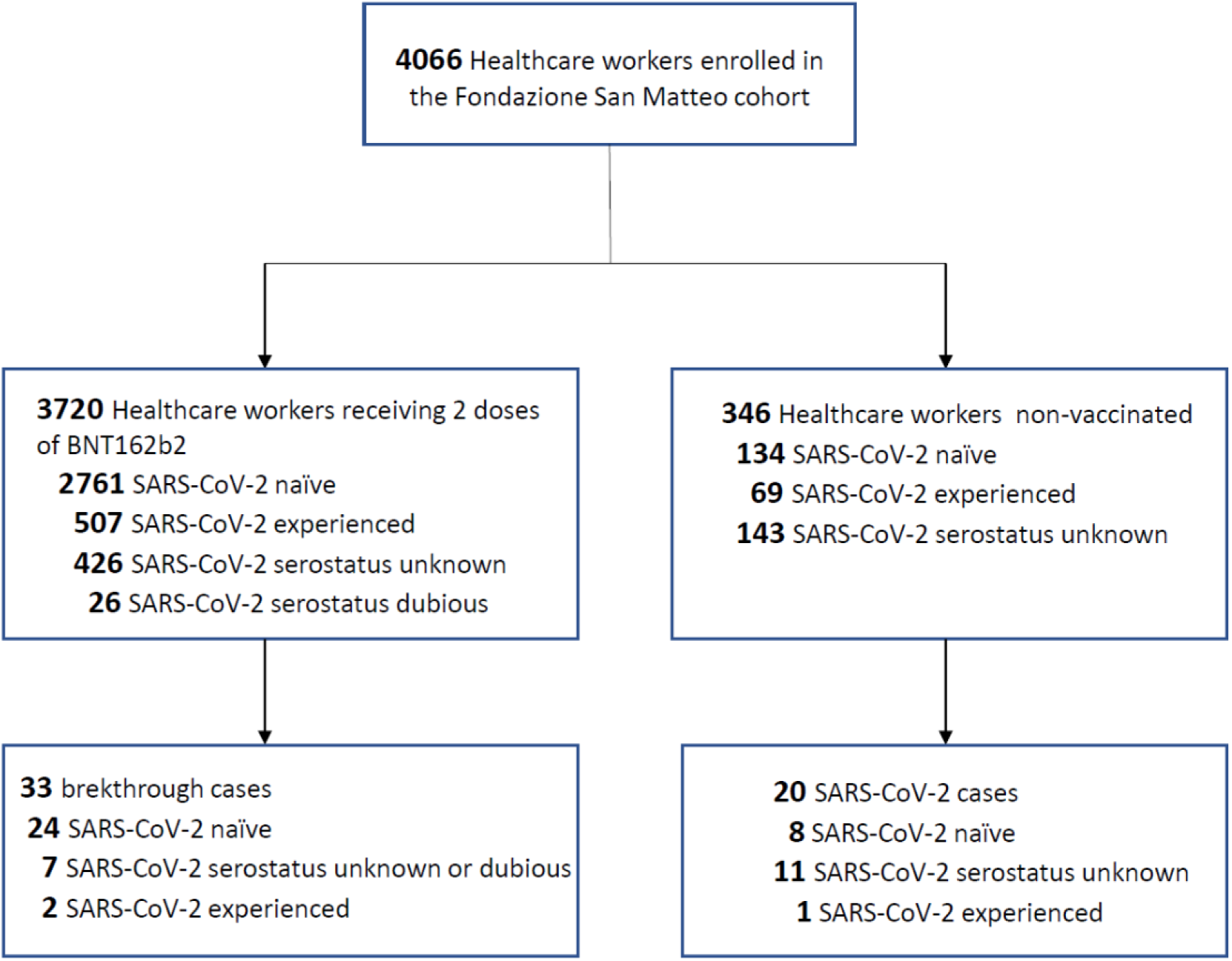
Study profile. Data from 4066 healthcare workers of San Matteo hospital, Pavia, Italy were analysed: 3720 subjects received complete schedule of BNT162b2 vaccine, while 346 subjects who did not receive vaccination during the study period were used as controls. According to serological or virological data subjects were considered SARS-CoV-2 experienced or naïve before initiation of the study, while SARS-CoV-2 serostatus was unknown or dubious for some individuals. SARS-CoV-2 infection was diagnosed during the study in 33 vaccinated subjects and 20 controls.

Of the 33 infected subjects, 23 (70%) were nursing or health care assistants, 8 (24%) were physicians, 1 (3%) was a healthcare technician and 1 (3%) was a researcher. The infection was symptomatic in 16 (48%) and asymptomatic in 17 (52%) subjects (Table 1). The most common symptoms reported were rhinitis in 9 (27%), cough in 3 (9%) and arthralgia in 3 (9%) infected subjects; more details on symptoms are described in Table 1. No subjects required hospitalization. Virus isolation from nasal swab was attempted in 21 subjects (13 symptomatic and 8 asymptomatic subjects). Infectious virus was recovered in 7/13 (54%) symptomatic and 4/8 (50%) asymptomatic subjects. Lineage characterization was available in 23 subjects in whom the amount of viral RNA was sufficient for genome sequencing. All analyzed patients were infected by the variant, also recently renamed as alpha variant. Evidence of virus transmission to family members or close contacts of the 33 infected subjects was observed in 2 (6.1%, 95% CI: 1.1-19.6%) cases, both of whom had a symptomatic infection.

**Table 1.** Symptoms of SARS CoV in vaccinated and non-vaccinated subjects Fisher’s Exact test (two-sided) was used for statistical analysis.

| Symptoms | Vaccinated subjects (n=33) | Non-vaccinated controls (n=20) | p-value |
| --- | --- | --- | --- |
| any | 16 (48%) | 17 (85%) | 0.010 |
| fever (>37.5°C) | 2 (6%) | 13 (65%) | <0.001 |
| headache | 2 (6%) | 2 (10%) | 0.627 |
| asthenia | 2 (6%) | 8 (40%) | 0.004 |
| arthralgia | 3 (9%) | 5 (25%) | 0.137 |
| pharyngodynia | 1 (3%) | 2 (10%) | 0.549 |
| rhinitis | 9 (27%) | 2 (12%) | 0.175 |
| cough | 3 (9%) | 3 (15%) | 0.661 |
| pneumonia | 0 (0%) | 2 (10%) | 0.138 |
| dyspnea | 0 (0%) | 4 (20%) | 0.017 |
| anosmia | 2 (6%) | 12 (60%) | <0.001 |
| ageusia | 1 (3%) | 10 (50%) | <0.001 |
| nausea | 1 (3%) | 2 (10%) | 0.549 |
| diarrhea | 1 (3%) | 2 (10%) | 0.549 |

The incidence of SARS-CoV-2 infection after vaccination was compared to that observed in the control group, including 346 healthcare workers of the same Institution that did not receive the vaccination during the study period (January-May 2021). The control group included 69 SARS-CoV-2 experienced and 134 SARS-CoV-2 naïve, while SARS-CoV-2 serostatus was unknown for 143 subjects (Fig 1). During January-May 2021, SARS-CoV-2 infection was detected in 20 non-vaccinated subjects: 1 subject among the 69 SARS-CoV-2-experienced, 8 among the 134 SARS-CoV-2 naïve individuals, and 11 among the 143 subjects with unknown serostatus (Fig 1). Of the 20 infected subjects, 12 (60%) were nursing or health care assistants, 4 (20%) were administrative workers, 2 (10%) were physicians, 1 (5%) was a healthcare technician and 1 (5%) was a maintenance worker. The infection was symptomatic in 17 (85%) and asymptomatic in 3 (15%) subjects, with a significantly higher occurrence of symptoms than in vaccinated subjects (p=0.010, Table 1). The most common symptoms reported were fever in 13 (65%), anosmia in 12 (60%), ageusia in 10 (50%) and asthenia in 8 (40%) infected subjects (Table 1), which were significantly more frequent than in vaccinated subjects (p≤0.004). Dyspnea, which occurred in 4 (8%) non-vaccinated controls, was also significantly higher in this group (p=0.017). Two subjects with pneumonia required hospitalization and one was transferred to Intensive Care Unit due to worsening of symptoms.

The 100-day cumulative incidence of SARS-CoV-2 infection in the overall population of vaccinated healthcare workers was 0.93% vs 5.78% (p<0.001) in the non-vaccinated control group (Fig 2a), with an hazard ratio (HR) of 0.17 (95%CI: 0.07-0.42%) and a protective effect in prevention of infection of 83% (95% CI: 58-93%). Excluding the 452 subjects with unknown or dubious serostatus, and considering separately experienced and naïve subjects (Fig 2b), the 100-day cumulative incidence was 0.42% in SARS-CoV-2-experienced and 0.90% in SARS-CoV-2 naïve subjects (p=0.272). Among non-vaccinated controls (Fig 2c), after exclusion of the 143 subjects with unknown serostatus the 100-day cumulative incidence was 1.45% in SARS-CoV-2-experienced and 5.97% in SARS-CoV-2 naïve subjects (p=0.139). The HR for developing SARS-CoV-2 infection in vaccinated vs non-vaccinated naïve subjects was 0.15 (95%CI: 0.03-0.76), with a protective effect of 86% (95% CI: 24-98%). The HR for developing secondary SARS-CoV-2 infection in vaccinated vs non-vaccinated naïve subjects was 0.29 (95%CI: 0.01-8.74), with a protective effect of 71% (95% CI: >0-99%). However, the number of non-vaccinated experienced subjects was too low (n=69) to detect a potentially significant effect in this latter comparison. The HR for developing SARS-CoV-2 infection after vaccination in experienced subjects vs non-vaccinated naïve subjects was 0.07 (95% CI: 0.01-0-31), with a protective effects of 93% (95% CI: 69-99%).

**Figure 2.**
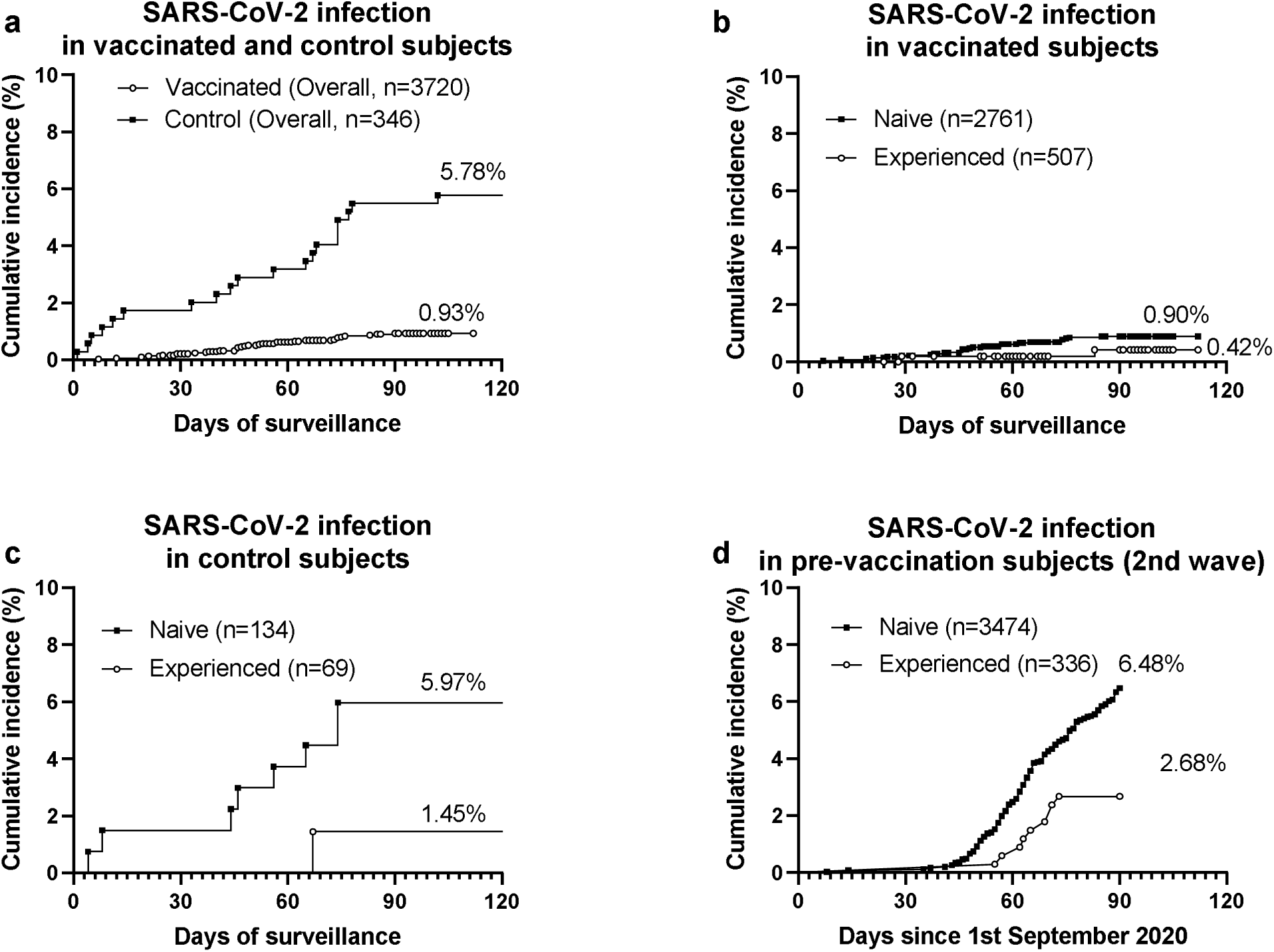
SARS-CoV-2 infection in vaccinated and non-vaccinated healthcare workers of San Matteo hospital. **a**, Cumulative incidence of SARS-CoV-2 infection in vaccinated subjects and non-Vaccinated controls. **b**, Cumulative incidence of SARS-CoV-2 infection in SARS-CoV-2-naïve and experienced vaccinated subjects. **c**, Cumulative incidence of SARS-CoV-2 infection in SARS-CoV-2-naïve and experienced non-vaccinated control subjects. **d**, Cumulative incidence of SARS-CoV-2 infection in SARS-CoV-2-naïve and experienced subjects before vaccine implementation during the second pandemic wave (period: September 1st-November 30th 2020).

### Protective effect of immunity elicited by natural SARS-CoV-2 infection against secondary infections

To compare the protective effect of the immunity elicited by vaccination or natural infection, we analyzed the incidence of SARS-CoV-2 infection in SARS-CoV-2-experienced or naïve healthcare workers from the same Institution during the second pandemic wave, before the implementation of the vaccination campaign. In the period April 29-June 30 2020, 3810 healthcare workers were tested for previous SARS-CoV-2 infection according to serostatus determination: 336 subjects resulted SARS-CoV-2 experienced and 3474 SARS-CoV-2 naïve. During the second pandemic wave, SARS-CoV-2 infection was detected in 9 SARS-CoV-2-experienced and 225 SARS-CoV-2 naïve subjects. The 3-months cumulative incidence of SARS-CoV-2 infection (Fig 2d) was 2.68% in experienced vs 6.48% in naïve subjects (p=0.006), with a hazard ratio of 0.41 (95%CI: 0.26-0.61). The protective effect of the immunity elicited by natural infection was 59% (95% CI: 39-74%) Data on symptoms were available for 112 subjects: 1/4 (25%) naïve and 85/108 (79%) experienced subjects developed upper respiratory symptoms and no patient required hospitalization.

### Immune response in vaccinated subjects with or without breakthrough SARS-CoV-2 infection

The antibody and T-cell response was determined in a subset of infected subjects within 48h after diagnosis of infection and in a control subset of non-infected subjects who were SARS-CoV-2 naïve before vaccination. Anti-S1/S2 IgG antibodies were detected in all but one of 27 infected subjects at levels not significantly different from that observed in 143 non-infected controls (Fig 3a). Neutralizing serum titer was determined in 24 infected subjects and compared to that observed in 128 controls (Fig 3b). Median serum neutralizing titer was 1:320 (range 1:10-≥1:640), slightly lower (two-fold) than that observed in controls (median titer: ≥1:640; range: 1:20-≥1:640; p=0.045). Finally, the T-cell response to peptide pool of the Spike protein was not significantly different between 24 infected subjects and 132 controls (Fig 3c). Antibody and T-cell response was not different between symptomatic and asymptomatic subjects (data not shown).

**Figure 3.**
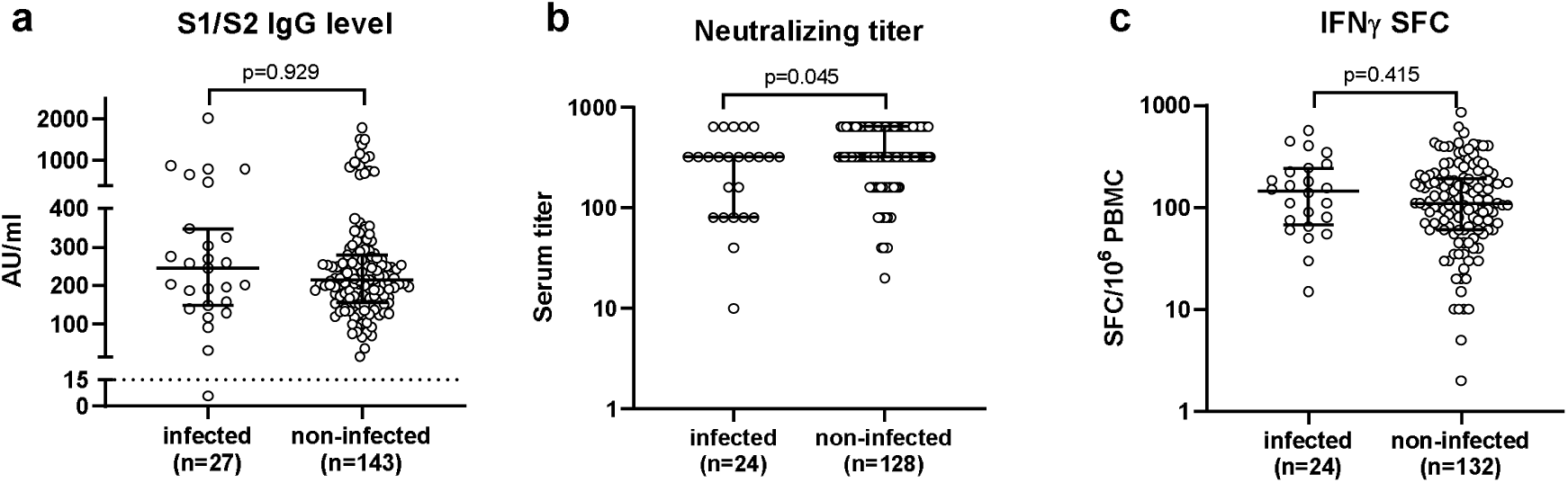
Antibody and T-cell responses in vaccinated healthcare workers with or without SARS-CoV-2 breakthrough infections. **a**, Anti-spike IgG level (AU: arbitrary units); **b**, serum neutralizing titer; **c**, IFNγ-producing spot forming cells (SFC)/10^6^ peripheral blood mononuclear cells (PBMC). Immune response was determined within 48h after diagnosis in infected subjects, and 21 days after complete vaccination schedule in non-infected subjects. Median values with interquartile ranges are shown. Mann-Whitney U-test (two-sided) was used for statistical analysis.

## Discussion

Results of this study suggest the protective effectiveness of BNT162b2 vaccine in prevention of SARS-CoV-2 infection in healthcare workers. By comparing the incidence of vaccine breakthrough infections with the incidence of SARS-CoV-2 infection in healthcare workers that did not receive the vaccination during the study period, an 83% protection from infection was calculated. All vaccine breakthrough infections were asymptomatic or symptomatic mainly with few and mild symptoms as rhinitis. The frequency of symptomatic infections was lower in vaccinated than non-vaccinated subjects (48% vs 85%). Live infectious virus was detected only in half of the cases of breakthrough infections and virus transmission to other individuals was documented in 6.1% of the cases. Finally, breakthrough infections were not associated with failure in developing antibody or T-cell response after vaccination.

The 83% effectiveness in prevention of infection of BNT162b2 vaccine observed in our study is slightly lower than the 95% efficacy reported in the phase 2/3 clinical trial [1] but is consistent with those reported in a retrospective analysis conducted in Israel [9] and in the SIREN study conducted in England [10], both involving healthcare workers. The active surveillance and screening performed in our as well as in the other above-mentioned studies may have led to the identification of a number of subclinical infections that were not ascertained in the clinical trial. Nevertheless, vaccine effectiveness observed by this real-world analysis in healthcare workers remains high, and results are consistent among different countries.

Protection from secondary infections after primary SARS-CoV-2 infection appears effective and sustained for at least 10 months [15]. The protection from subsequent SARS-CoV-2 infection provided by the vaccine appears even higher than that induced by natural infection (83% vs 59%, as observed in the retrospective analysis of our study). We analysed separately the incidence of breakthrough infections in individuals who were either SARS-CoV-2 naïve or experienced before vaccination. Although we observed a lower incidence of breakthrough infections in experienced than naïve subjects (0.42% vs 0.90%) and a higher protective effectiveness (94% vs 83%), the population size was too low to verify whether SARS-CoV-2-experienced achieve a significantly higher protection from infection after vaccination than naïve subjects. In addition, we cannot exclude that previously infected individuals may have had different behaviors or underlying risk profiles. Nevertheless, immunogenicity studies showed that vaccination is able to boost the pre-existing immunity in experienced subjects, who develop higher antibody and T-cell levels than naïve subjects [16–18]. In addition, we observed that vaccination of experienced individuals elicited neutralizing antibody at levels that overcome the partial antibody escape of the B.1.351 variant [18]. It is tempting to speculate that three antigenic exposures, as in the case of SARS-CoV-2-experienced subjects receiving two vaccine doses, may confer a higher and broader protection.

Breakthrough infections were asymptomatic or symptomatic with few symptoms. To assess whether the detection of SARS-CoV-2 RNA in naso-pharyngeal samples of vaccinated subjects with breakthrough infection was associated with the presence of live infectious virus, we attempted virus isolation on cell cultures in 21 of the 33 subjects, recovering infectious virus only in half of the cases. We could not compare the rates of infectious virus recovery in vaccinated vs unvaccinated SARS-CoV-2 RNA positive subjects, since we did not perform virus isolation in the control group. However, the low rate of detection of infectious virus is in line with a recent report showing decreased viral load in infected vaccinated subjects [19], and suggests a lower contagiousness, along with the lower severity, of the vaccine breakthrough infections. Thus, SARS-CoV-2 RNA detected in fully vaccinated is likely to be often a sign of an abortive infection limited and blocked on mucosal surfaces by the elicited immunity.

Most importantly, we documented a lack of transmission of the virus in the great majority of cases, since only 2/33 subjects transmitted the infection to a family member. Transmission was ascertained by contact tracing and analysis of naso-pharyngeal swabs of coworkers and family members. Although we cannot exclude that the actual rate of transmission could have been underestimated, this data suggests a low contagiousness of SARS-CoV-2 infection in vaccinated subjects and the effectiveness of SARS-CoV-2 vaccination in blocking the infection spreading at the population level. However, this observation should be confirmed in controlled studies.

In order to verify the hypothesis that a poor individual response to the vaccine is the cause of breakthrough infections, we compared the antibody and T-cell responses of the infected individuals with a control group of vaccinated uninfected subjects. Since the immune response was evaluated in the peri-infection period (within 48h after diagnosis), it is unlikely that the levels of antibody and T cells observed are the consequence of a boost due to the infection. No difference was observed for anti-Spike IgG and T-cells, while a slight reduction in the neutralizing serum titers was detected in the infected subjects. However, it was not possible to define a cutoff level of neutralizing titer able to identify poor responders that are at higher risk for vaccine breakthrough infections. We could exclude also that a viral variant associated to potential vaccine escape was causing the infection, since in all cases in which the RNA amount was sufficient for sequencing, the B.1.1.7 variant (alpha), which was accounting for the great majority of viral strains circulating in Italy during the study period, was detected. Before the end of the study, in Italy the first cases of delta variant have been reported, accounting for 0.02% [20]. No cases of delta variant were identified in vaccine breakthrough infections of our study.

The strength of this study resides in the prospective systematical collection of virological data (viral RNA detection and infectious virus recovery), clinical symptoms, immune response and virus transmission to other individuals, providing an insight on the characteristics of vaccine breakthrough infection and initial observational evidence for their low contagiousness. The limitation resides in the small sample size of the control group for the evaluation of vaccine effectiveness, and the lack of a control for the analysis of the contagiousness of breakthrough infections. Nevertheless, the observed vaccine effectiveness was similar to that observed in other real-world analysis on healthcare workers. Notwithstanding the non-random nature of the control group, we can assume that both vaccinated and control subjects were exposed to the same risk of SARS-CoV-2 infection, since all of them were healthcare workers of the same Institution. The decision to defer vaccination was taken by the individual subjects.

In conclusion, our analysis support the effectiveness of the BNT162b2 vaccine in preventing SARS-CoV-2 infection, and suggest that breakthrough infections are poorly symptomatic and likely associated with a low contagiousness. Nevertheless, the duration of the vaccine protection should be evaluated in the next future.

## Methods

### Study subjects and design

The occurrence of SARS-CoV-2 infection was monitored prospectively in 3720 healthcare workers of Fondazione IRCCS Policlinico San Matteo, Pavia receiving 2 doses of the BNT162b2 vaccine. Subjects completed vaccination schedule between January 18 and March 31, 2021 and data were collected until May 10, 2021.

Data on SARS-CoV-2 infection in 346 healthcare workers of the same Institution that did not receive the vaccination during the study period were used as control (data were collected in the period January 1-May 10, 2021).

Data on incidence of SARS-CoV-2 infection during the second pandemic wave in 3810 healthcare workers from the same institution who had serological definition of previous SARS-CoV-2 infection were used to compare protection from SARS-CoV-2 infection induced by the vaccine or by natural infection (data were collected in the period September 1^st^-November 30, 2020). Subjects analysed are a subgroup of a cohort of healthcare workers partially described in [21].

Naso-pharyngeal swabs were collected and tested for SARS-CoV-2 RNA positivity in subjects with symptoms suggestive for SARS-CoV-2 infection or in case of contact with infected subjects as previously reported [22]. In vaccinated subjects who resulted SARS-CoV-2 RNA-positive, blood samples for immune response analyses were collected 24-48h after nasopharyngeal swab sampling. The transmission of the infection from SARS-CoV-2 RNA-positive vaccinated subjects was investigated by contact-tracing and monitoring for SARS-CoV-2 RNA detection in nasal swabs in coworkers and family members of the infected subjects. Data on symptoms were collected during an interview by a physician and inserted into a specific database. Study procedures were approved by Fondazione IRCCS Policlinico San Matteo and Comitato Etico Area Pavia and all the subjects gave their written informed consent.

### Virus isolation

Virus isolation was performed by inoculation of 200μl nasopharyngeal swab suspension medium, after decontamination with an antibiotics pool for 30 min at room temperature, on Vero E6 cells cultured in 24well plate, and detection of cytopathic effect after one-week culture.

### Sequencing

Lineage characterization was available in 23 subjects in whom the amount of viral RNA was sufficient for genome sequencing. In detail, total RNAs were extracted from nasopharyngeal swabs by using QIAamp Viral RNA Mini Kit (Qiagen, Hilden, Germany), followed by purification with Agencourt RNA Clean XP beads (Beckman Coulter, Inc., Brea, CA). Both the concentration and the quality of all RNA samples were measured and checked with the Nanodrop. Virus genomes were generated by using a multiplex approach, using version 1 of the CleanPlex SARS-CoV-2 Research and Surveillance Panel (Paragon Genomics, Hayward, USA), according to the manufacturer’s protocol starting with 50⍰ng of total RNA. Briefly, the multiplex PCR was performed with two pooled primer mixtures and the cDNA reverse transcribed with random primers was used as a template. After ten rounds of amplification, the two PCR products were pooled and purified. Then the digestion reaction was performed to remove non-specific PCR products, followed by second PCR reaction for barcoding with 24 rounds of amplification. Libraries were checked using High Sensitivity Labchip and quantified with Qubit Fluorometric Quantitation system (Life Technologies, Carlsbad, CA, USA). Equimolar quantity of libraries was pooled, and the obtained run library mix was loaded at 1.5⍰pM into Illimuna MiSeq platform (Illumina, San Diego, CA, USA,) for sequencing [23]. NGS data were also analysed with an in-house pipeline and lineages were assigned from alignment file using the Phylogenetic Assignment of Named Global Outbreak LINeages tool PANGOLIN v1.07 (https://github.com/hCoV-2019/pangolin).

### SARS-CoV-2-specific antibody and T-cell determination

To detect subjects with SARS-CoV-2 infection after the first pandemic wave, serological analysis was performed in the period April 29-June 30 2020 using chemiluminescent assay (Liason SARS-CoV-2 S1/S2 IgG, Diasorin, Saluggia, Italy) for the quantitative measurement of SARS-CoV-2 anti-S1 and anti-S2 IgG antibody. Results higher than 15 AU/mL were considered positive and defined SARS-CoV-2 experienced subjects, whereas results below 12 AU/mL were considered negative and defined SARS-CoV-2 naïve subjects. Subjects with borderline results ranging from 12 and 15 AU/mL were not included in the analysis. A further serological screening was conducted after the second pandemic wave in the period December 15 2020-February 3 2021. Since this second screening overlapped with the initiation of the vaccination campaign, the electrochemiluminescent assay Elecsys Anti-SARS-CoV-2 N (Roche Diagnostics Rotkreuz, Switzerland), which provides quantitative measures of mainly IgG (but also IgA and IgM) specific for SARS-CoV-2 Nucleocapsid protein (not present in the vaccine) was used. Results were given as units (U)/ml and are considered positive when ≥0.8 U/ml. Subjects with positive serological results after either or both screenings were considered SARS-CoV-2 experienced before vaccination.

Antibody and T-cell response to Spike protein was determined in vaccinated subjects after detection of SARS-CoV-2 infection and in a control group of SARS-CoV-2-naïve subjects 21 days after complete vaccination [18]. Anti-Spike IgG antibody was determined using Liason SARS-CoV-2 S1/S2 IgG (Diasorin). Neutralizing antibody serum titre was determined as following described [24]. Fifty μl of serum, from 1:10 to 1:640 in a serial fourfold dilution, were added in two wells of a flat bottom tissue culture microtiter plate (COSTAR, Corning Incorporated, NY 14831, USA). Then, the same volume of 100 TCID50 of SARS-CoV-2 strain was added and plates were incubated at 33°C in 5% CO_2_. All the dilutions were made in E-MEM with addition of 1% penicillin, streptomycin and glutammin and 5 γ/mL of trypsin. After 1 h incubation at 33°C 5% CO_2_, Vero E6 cells were added to each well. After 72 h of incubation at 33°C 5% CO_2_, plates were stained with Gram’s crystal violet solution (Merck KGaA, 64271 Damstadt, Germany) plus 5% formaldehyde 40% m/v (Carlo ErbaSpA, Arese [MI], Italy) for 30 min. Microtiter plates were then washed under running water. Wells were scored to evaluate the degree of cytopathic effect (CPE) compared to the virus control. Blue staining of wells indicated the presence of neutralizing antibodies. Neutralizing titer was the maximum dilution with the reduction of 90% of CPE. Results higher or equal to 1:10 serum titer were considered positive.

Spike specific T-cell response was determined with an IFNγ ELISpot assay after peripheral blood mononuclear cells (PBMC) stimulation with a peptide pool (15mers overlapping by 10 aminoacids) spanning the entire Spike protein [25]. In more details, PBMC were isolated from heparin-treated blood by standard density gradient centrifugation. The number of IFNγ-producing spot forming cells (SFC) was determined by ELISpot. Briefly, PBMC (2×10^5^/100μl culture medium per well) were stimulated in duplicate for 24 h in 96-well plates (coated with anti-IFN-γ monoclonal capture antibody) with peptide pools (15 mers, overlapping by 10 aminoacids, Pepscan, Lelystad The Netherlands) representative of the S at the final concentration of 0.25 μg/ml. Phytoheamagglutinin (PHA; 5 μg/mL) was used as positive control, and medium alone as negative control. After washing, plates were incubated for 90 minutes at 37°C with biotinylated IFN-γ detection antibody. Then, streptavidin-alkaline phosphatase conjugate was added, and plates were incubated at 37°C in a 5% CO_2_ atmosphere for 1 hour. Finally, and 5-bromo-4-chloro-3-indolyl phosphate/nitro blue tetrazolium (BCIP/NBT) was added for 20 minutes at room temperature. Wells were then washed several times under running water and air-dried overnight. Spots were counted using an automated AID ELISPOT reader system (AutoImmun Diagnostika GmbH, Strasburg, Germany). The mean number of spots from duplicate cultures were adjusted to 1 × 10^6^ PBMC. The net spots per million PBMC was calculated by subtracting the number of spots in response to negative control from the number of spots in response to the S or N antigen. Responses ≥10 net spots/million PBMC were considered positive based on background results obtained with negative control (mean SFC+2SD).

### Statistical analysis

The cumulative incidence of SARS-CoV-2 infection was calculated with the Kaplan-Meier method and log-rank test was adopted for statistical comparison. The hazard ratio (HR) and 95% confidence interval (CI) for SARS-CoV-2 infection in the various groups of patients were calculated with the log-rank approach. The vaccine effectiveness in prevention of SARS-CoV-2 infection was calculated as 100% x (1-HR), and the relevant 95% CI were calculated as 100% x (1-95% CI_HR_), where CI_HR_ is the 95% CI of the HR. Similarly, the protective effectiveness of immunity elicited by SARS-CoV-2 infection against secondary infections was calculated as 100% x (1-HR), and the relevant 95% CI were calculated as 100% x (1-95% CI_HR_). The frequency of symptoms in vaccinated and control subjects were compared with the Fisher’s exact test. The antibody and T-cell levels in vaccinated subjects with or without breakthrough infections were compared with the Mann-Whitney U-test.

## Data Availability

The data that support the findings of this study are available from the corresponding author upon reasonable request.

## Data availability

The anonymized data relevant to SARS-CoV-2 infection during the study period in vaccinated and control subjects, along with serologic results indicating their previous exposure to SARS-CoV-2 infection, are available in the Dryad database under DOI 10.5061/dryad.n2z34tmxk [https://doi.org/10.5061/dryad.n2z34tmxk]. Raw data associated to Figures 2a-c are present in the dataset. The 23 SARS-CoV-2 sequences obtained in this study are openly available on GISAID portal and European Nucleotide Archive under the accession numbers EPI_ISL_3836237-EPI_ISL_3836259.

## Acknowledgments

This work was supported by Fondazione Cariplo [grant CoVIM, no. 2020-1374] and Ministero della Salute, Ricerca Finalizzata [grants no. COVID-2020-12371760 and COVID-2020-12371817]. AP and FB have received funding from the European Union’s Horizon 2020 research and innovation programme under grant agreement No 101003650.

## Author contribution

FR and DL analyzed and interpreted the data and drafted the manuscript; AM, VN, AM, AMG, GG, collected and managed the data; CR, MD performed patients’ follow-up; FeB and IC, performed experiments on T-cell response; JCS, AF, AS performed experiments on antibody response; EP performed virus isolation; SP, AP, FG sequenced viral genomes; CM, AT supervised participants enrollment; FaB supervised the study and revised the manuscript. All the authors critically reviewed the manuscripts.

